# Cracking the Code: A Scoping Review to Unite Disciplines in Tackling Legal Issues in Health Artificial Intelligence

**DOI:** 10.1101/2024.04.22.24305806

**Authors:** Sophie Nunnelley, Colleen M. Flood, Michael Da Silva, Tanya Horsley, Sarathy Kanathasan, Bryan Thomas, Emily Ann Da Silva, Valentina Ly, Ryan C. Daniel, Mohsen Sheikh Hassani, Devin Singh

## Abstract

**Objectives:** The rapid integration of artificial intelligence (AI) in healthcare requires robust legal safeguards to ensure safety, privacy, and non-discrimination, crucial for maintaining trust. Yet, unaddressed differences in disciplinary perspectives and priorities risks impeding effective reform. This study uncovers convergences and divergences in disciplinary comprehension, prioritization, and proposed solutions to legal issues with health-AI, providing law and policymaking guidance.

**Methods:** Employing a scoping review methodology, we searched MEDLINE® (Ovid), EMBASE (Ovid), HeinOnline Law Journal Library, Index to Foreign Legal Periodicals (HeinOnline), Index to Legal Periodicals and Books (EBSCOhost), Web of Science (Core Collection), Scopus, and IEEE Xplore, identifying legal issue discussions published, in English or French, from January 2012 to July 2021. Of 18,168 screened studies, 432 were included for data extraction and analysis. We mapped the legal concerns and solutions discussed by authors in medicine, law, nursing, pharmacy, other healthcare professions, public health, computer science, and engineering, revealing where they agree and disagree in their understanding, prioritization, and response to legal concerns.

**Results:** Critical disciplinary differences were evident in both the frequency and nature of discussions of legal issues and potential solutions. Notably, innovators in computer science and engineering exhibited minimal engagement with legal issues. Authors in law and medicine frequently contributed but prioritized different legal issues and proposed different solutions.

**Discussion and Conclusion:** Differing perspectives regarding law reform priorities and solutions jeopardize the progress of health-AI development. We need inclusive, interdisciplinary dialogues concerning the risks and trade-offs associated with various solutions to ensure optimal law and policy reform.

**KEY MESSAGES:** *What is already known on this topic:* There has been no systematic examination of the multidisciplinary literature discussing legal challenges posed by health-AI. Prior efforts have addressed ethical concerns or limited subsets of legal issues or technologies, and therefore do not establish the comprehensive groundwork essential for fostering meaningful cross-disciplinary dialogue on health-AI regulation.

*What this study adds:* Our study uncovers a shared interdisciplinary apprehension regarding the effective regulation of health-AI. However, distinct stakeholders such as physicians, innovators, and legal scholars hold divergent perspectives on these issues and their relative significance. Notably, certain critical voices, such as within discussions around informed consent, are conspicuously absent, hindering the prospects of effective reform.

*How this study might affect research, practice, or policy:* The findings underscore the imperative for governments to facilitate inclusive dialogue and reconcile disparate disciplinary viewpoints. Effective regulation is pivotal in ensuring the safe and responsible deployment of health-AI for the public good. This study presents essential entry points for the much-needed discourse on this challenge facing governments around the world.

## I. INTRODUCTION

Artificial intelligence (AI) is transforming healthcare with the promise of more accurate diagnoses, improved treatment options, and a restoration of humanized care through the automation of administrative tasks. [1][2] But a roadblock is uncertainty about how to manage its risks, for instance, relating to patient privacy, blurred responsibility for mistakes made by AI, and the potential for patient harm from algorithmic bias. There is growing recognition of the urgent need for regulation to ensure health-AI is developed in a responsible manner. [3–10] This need is amplified by the current generative AI arms race between behemoth technology companies like Open AI, Microsoft, and Google, and the active integration of these tools into healthcare delivery. [11, 12]

But what is the pathway to effective law reform? Many agree that effective reforms will require “multidisciplinary, international effort.” [5] Yet, disciplines too-often talk only to one another, impeding the joint-conversations and analyses that are essential for both understanding the nature of the problem (e.g. what risks do generative AI models pose to privacy?) and how to resolve them. Addressing the urgent need for cross-disciplinary understanding, we provide a first-of-its-kind systematic examination of which legal concerns are raised and how they are discussed by different disciplines. We find a shared concern for better health-AI regulation. Yet, understandings of key legal issues and solutions remain fractured. Multidisciplinary work is essential to ensure law reform incorporates a genuine understanding of AI, including its effects on patients and the clinicians tasked with employing AI at the bedside.

## II. METHODS

Over the last decade, the health-AI literature has surged from a trickle to a torrent. Employing a scoping review, we systematically mapped the legal concerns about health-AI raised in the published literature by different disciplines, including medicine, law, nursing, pharmacy, other healthcare professions (dentistry, nutrition, etc.), public health, computer science, and engineering. We aimed to assess which legal concerns were raised, how they were characterized, and what solutions were proposed by these disciplines. [13] Our review was guided by an a priori protocol and conducted in accordance with the Arksey and O’Malley framework as extended by Levac et al. [13–15] Reporting was informed by the PRISMA Extension for Scoping Reviews (PRISMA-ScR), including the six Arskey and O’Malley stages. [14]

### Stage 1: Identifying the Research Question(s)

The primary research question was (1) What is known from the literature regarding legal concerns in health-related AI? Secondary questions were (2) Are the legal concerns identified explicitly prioritized? and (3) Do different disciplines identify, represent, or prioritize legal concerns differently?

### Stage 2: Search Strategy and Selection Criteria

Guided by two trained librarians, a preliminary search of MEDLINE® and HeinOnline was conducted to pilot test a highly sensitive search strategy for its ability to identify key articles. Refinements led to a final MEDLINE® search strategy, which was peer reviewed using the Peer Review of Electronic Search Strategies checklist and adapted to other databases. [15]

The following electronic databases were searched on July 21, 2021 for eligible records published on or after January 1, 2012: MEDLINE® (Ovid), EMBASE (Ovid), HeinOnline Law Journal Library, Index to Foreign Legal Periodicals (HeinOnline), Index to Legal Periodicals and Books (EBSCOhost), Web of Science (Core Collection), Scopus, and IEEE Xplore (supplemental e-table 1). For full search terms (used for MEDLINE® and adapted to other databases) see supplemental e-table 2 or the Protocol. [13] Searches were augmented by hand-searching reference lists of relevant full-text records. [16] All records were imported into a proprietary review software program (Covidence®) for duplicates removal and eligibility assessment.

### Stage 3: Study Selection

All English and French-language records discussing legal concerns or solutions regarding health-AI were selected. For definitions of “legal concern”, “artificial intelligence” and “health-related”, see supplemental e-table 3. We excluded records raising issues that were characterized solely in ethical terms, without legal import or analysis, and abstracts and conference proceedings and secondary syntheses. Systematic reviews were tracked to ensure inclusion of relevant primary sources. [13]

Decisions regarding record inclusion were made by two authors with guidance from a pilot-tested eligibility assessment form and using record management software. Agreement was assessed and reported using a Kappa statistic. [17] Subject matter expert authors resolved any conflicting decisions. Of 18,168 identified records, 432 studies were included for analysis. A summary of inclusion decisions at each stage is provided in the PRISMA flow diagram (Figure 1).

**Figure 1:**
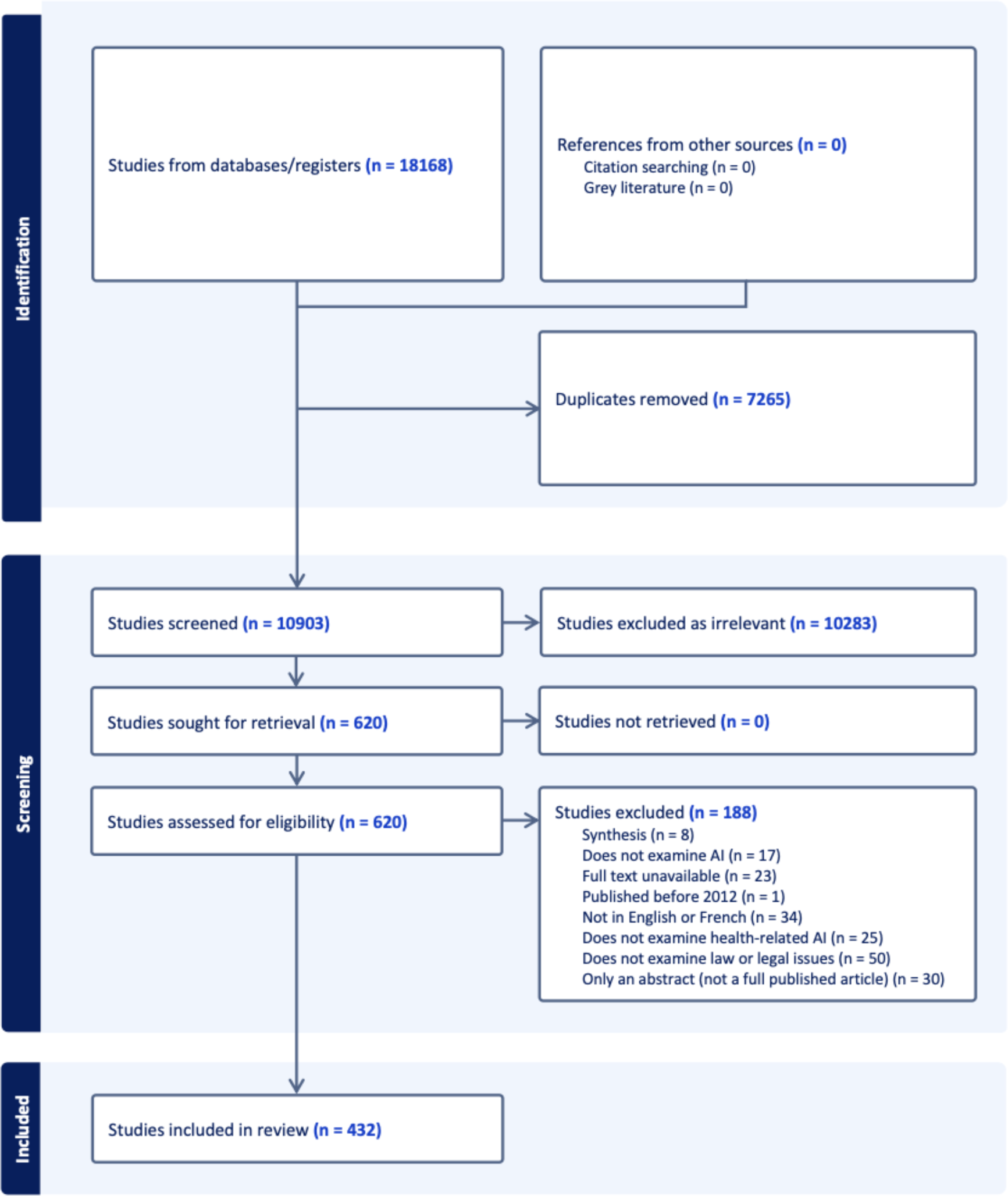
PRISMA Diagram.

### Stage Four: Extracting and Charting the Data

We developed, pilot tested and refined a standardized data extraction tool until it was deemed to support data extraction at a high-level of consistency. Law students under author supervision extracted (1) record-level demographic information and (2) text-based expressions of legal concerns, express prioritizations, and proposed solutions. Information was extracted verbatim without any attempt at interpretation. Extracted demographic information included the faculty of the corresponding author, which was deemed the author’s “discipline” for analysis purposes. (Supplemental e-table 4).

Discussions of legal issues were extracted using a list of ten legal issues and an ‘other’ category. Where an issue could be categorized under two headings (e.g., data leaks could be described as a privacy or cybersecurity issue), extraction followed the characterization in the text. For issues that could be discussed in a legal or non-legal way (e.g., safety), extraction was only done if the issue was discussed as a legal issue. Where an article proposed law reform, regulatory, or other solutions to one or more of the problems it identified, the data was extracted and categorized into solution type (supplemental e-table 4).

### Stage 5: Collating, Summarizing, and Reporting Results

Quantitative data was extracted, analyzed, and visually represented with the aid of custom software written in Python. To generate qualitative data, we collated the extracted data by legal concern or solution type, further stratifying it by author discipline. We then closely reviewed to identify themes, and prepared summary analyses for each legal issue, identifying key similarities and differences between disciplines. Issue coding and summary analyses were reviewed and confirmed by a second author.

### Stage 6: Stakeholder Consultation

In March 2023 we conducted a consultation process with an International Advisory Board composed of multi-disciplinary experts. [18] Members reviewed the face validity of initial findings and confirmed that our results align with their understanding of the legal landscape of health-AI.

### Patient and Public Involvement

The International Advisory Board for the project includes a member who is a patient partner, caregiver, and advocate for the co-design of research and healthcare. We consulted with this member during our Stakeholder Consultation.

## III. RESULTS

### What is the frequency and distribution of legal issues discussions?

We found exponential growth in the literature raising legal issues with health-AI. Rates of discussion grew by 950% between 2012-2016 and by 2914% between 2016 and 2020 (Figure 2). The geographic distribution of published legal concerns was USA-led (38%), followed by the UK (9%), Canada (7%), and Australia (5%). Many countries were marginally represented or unrepresented (Figure 3). Authors raising legal issues most frequently were in medicine (36%) followed by law (28%) (Figure 4). AI developers (represented by authors in computer science and engineering) were minimally represented in the literature, with 4% for each of those disciplines.

**Figure 2:**
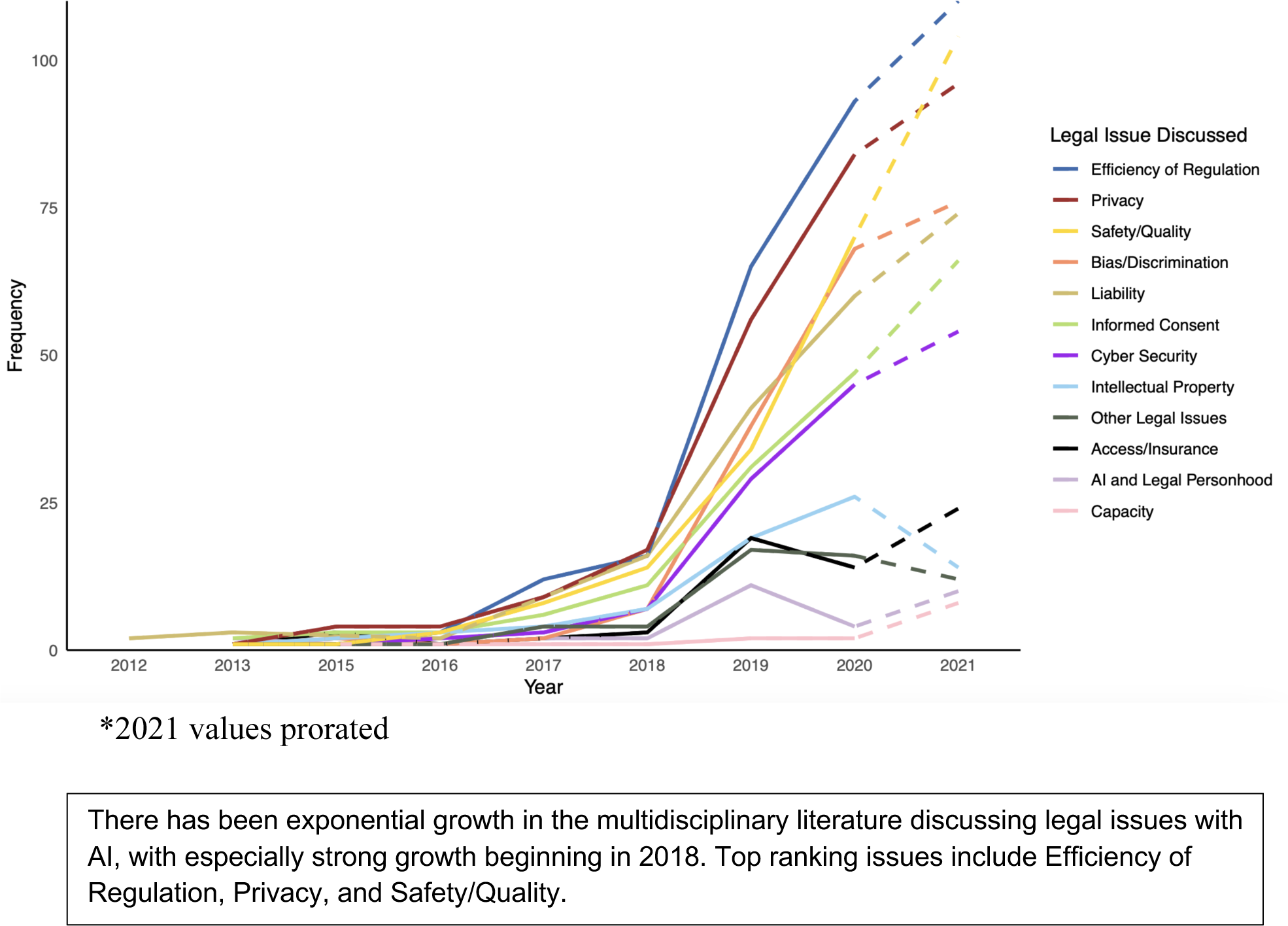
Growth in discussions of legal issues with health-AI from 2012 to 2021*.

**Figure 3:**
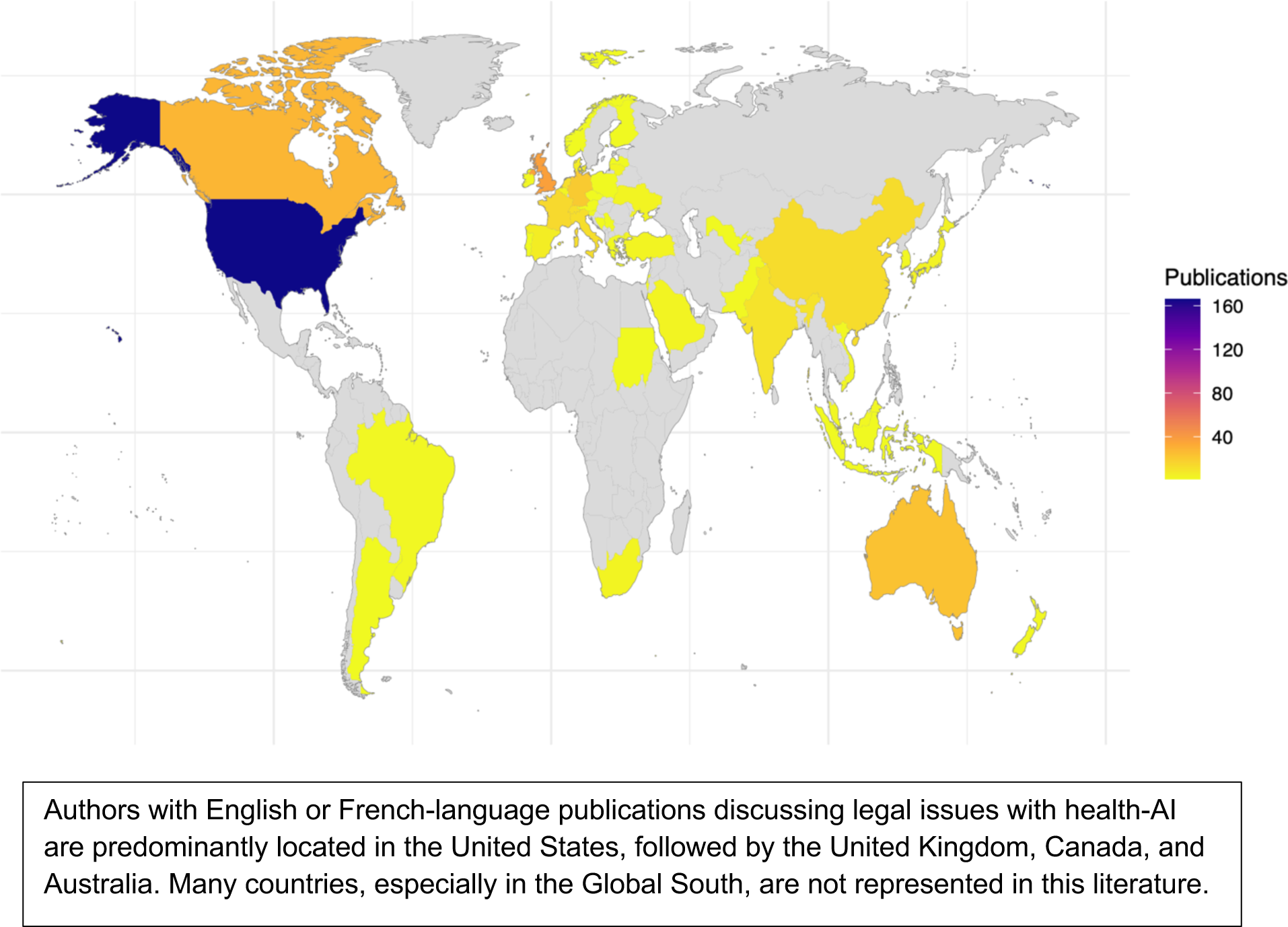
Geographical distribution of publications discussing legal issues with health AI (2012-2021)

**Figure 4:**
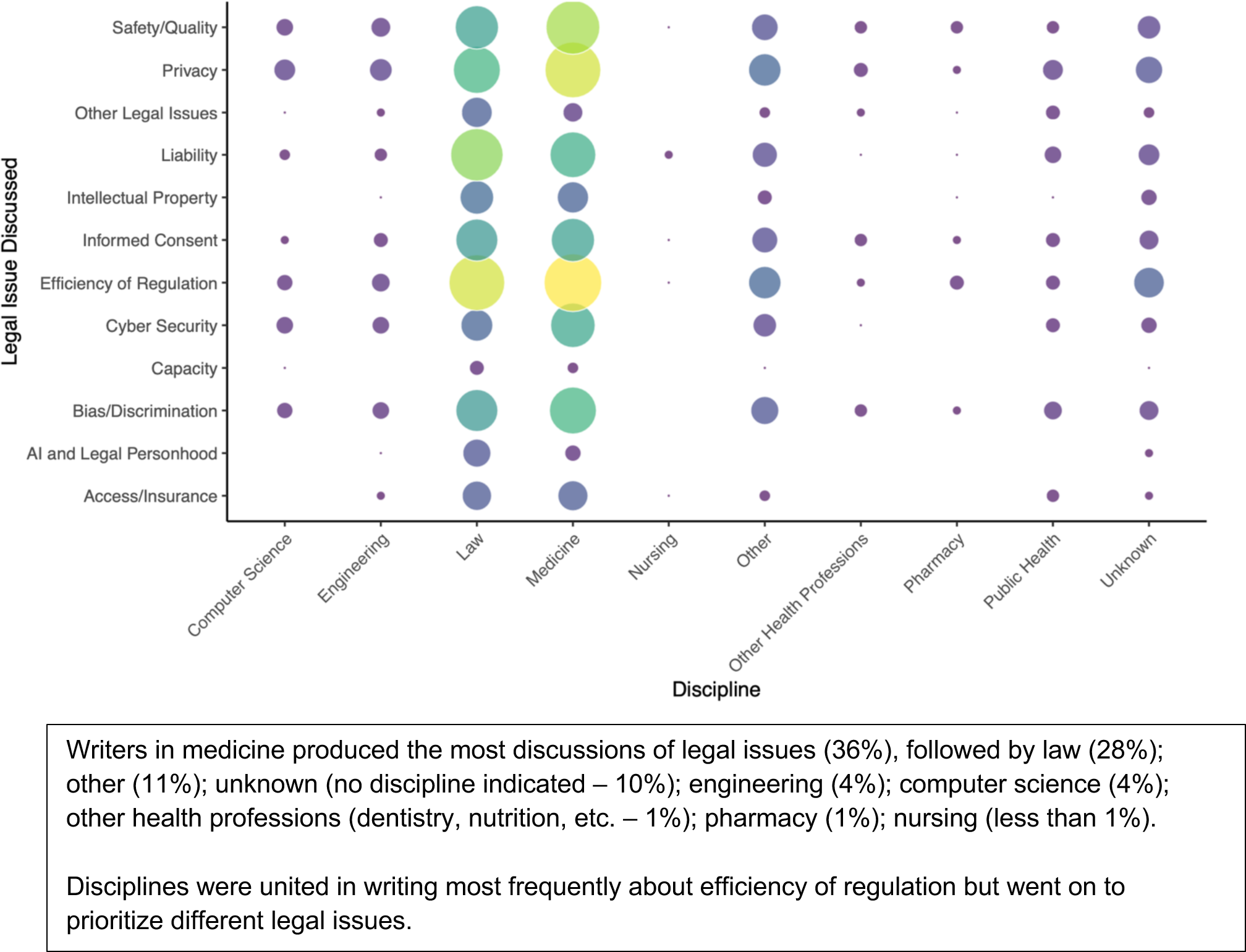
Disciplinary distributions of discussions of legal issues with health-AI (2012-2021)

Overall, the most commonly discussed legal issue was a concern to ensure the efficiency of regulation – for instance, the worry that unclear, overzealous, or inconsistent regulation will make compliance difficult or impede innovation. Concerns over regulatory efficiency accounted for 17% of legal issues discussions, and this issue ranked first for authors in each of medicine and law. After this issue, authors in medicine most often discussed privacy, followed by safety/quality, while legal authors more often discussed liability, followed by privacy.

The most frequently discussed solutions to legal issues were new legislation (28%) and voluntary improvements (i.e., non-legal measures; 26%), with calls to reform existing laws comprising 14% of solutions discussions. Authors in medicine and law again dominated these discussions (Figure 5). Authors in medicine were most likely to discuss voluntary improvements (33%), followed by new legislation (23%). They also discussed other non-legal instruments for promoting responsible health-AI adoption (e.g., mandatory training and professional guidelines) more frequently than legal writers. Legal authors more often discussed new legislation (34%; such as dedicated AI legislation) [19], followed by reform of existing law (22%; for instance, to strengthen privacy protections).

**Figure 5:**
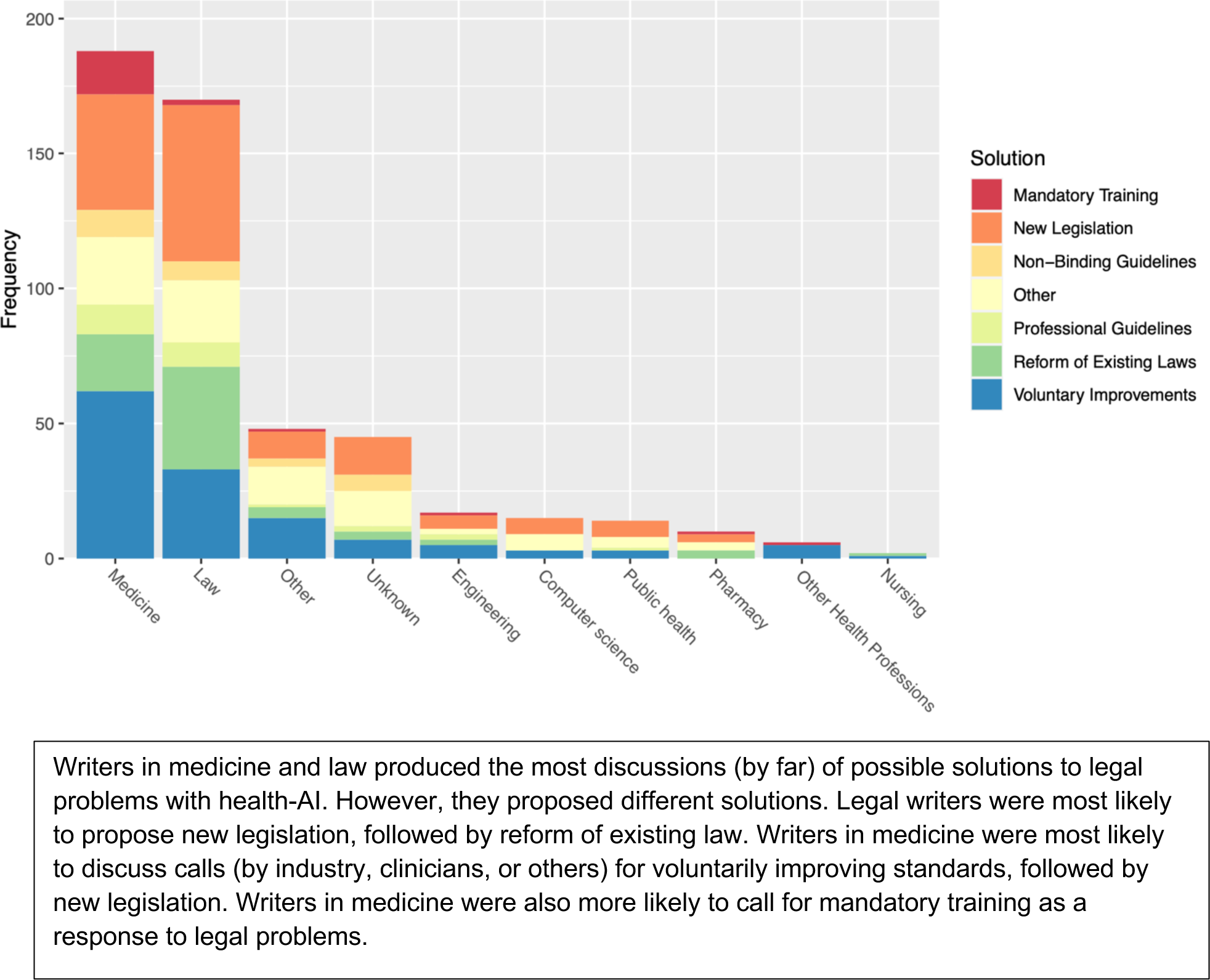
Disciplinary distribution of references to solutions to legal issues with health-AI (2012-2021)

### How do different disciplines characterize and prioritize legal issues?

We identified themes in how disciplines represent legal issues, noting similarities and differences across disciplines (see supplemental e-table 5). On some issues we found significant cross-disciplinary agreement, including:

- The need for efficient regulation and consensus that existing safety and quality regulations are inadequate, inconsistent, or otherwise not ‘fit for purpose’ for health-AI.
- The lack of clarity as to who, as between developers, healthcare institutions, or clinicians, should bear liability when AI use results in patient harm. (As put in one paper, “all parties face an uncertain liability landscape” [20]).
- The critical need for improved cybersecurity where health-AI is employed.
- The importance of addressing the risk of algorithmic bias, which can pose safety risks to patients and exacerbate existing inequities.

On other legal issues we found distinct disciplinary characterizations and approaches. For example:

- While few authors overall discussed whether AI-use must be disclosed to the patient as part of informed consent to treatment, those who discussed the issue were usually in law. [21] When writers in medicine discussed “consent”, they usually were referring to the collection, use, and disclosure of personal information – that is, consent as it relates to the exercise of patient privacy rights.
- Disciplines made different predictions about the likely allocation of liability where health-AI use results in patient harm. Legal authors more often predicted reduced physician liability due to greater overall accuracy and the eventual incorporation of AI into the standard of care, while writers in medicine worried about possible increased risk that clinical reliance on AI will be deemed negligent. [22–26]
- Authors from different disciplines emphasized varying concerns about access and equity. For instance, legal writers were more likely to note risks from third parties like insurers, who may refuse to cover AI-based care or refuse to cover harms relating to AI use, or who may use AI-tools to deny coverage for healthcare more broadly, leading to inequity. Authors in medicine more frequently raised a possible “digital divide” between healthcare institutions and associated patient populations that are able or unable to afford AI-based medicine.

## IV. DISCUSSION

### Overview

AI holds the potential to revolutionize healthcare by ushering in a new era of precision medicine and alleviating the strain on overburdened healthcare systems. However, there is a growing consensus that realizing AI’s potential requires adequate legal governance. Given the rapid evolution of AI technology, delivering optimal regulation presents a significant challenge. Addressing this challenge necessitates convergence across disciplines to identify the specific risks posed by AI in healthcare and determine the best approaches to regulation.

The situation resembles the tale of The Blind Men and the Elephant, where each discipline perceives only a fragment of the intersecting issues, hindering a comprehensive understanding. Regulatory efforts based on incomplete disciplinary perspectives risk distortion or failure. For instance, the Canadian government’s introduction of the Artificial Intelligence and Data Act in 2022 faced criticism for its vagueness, prompting calls for stakeholder consultation and consensus-building. [27–28] This study supports the necessary conversations by showing us who is most discussing different legal risks, which voices are missing, and how disciplines characterize the risks and possible solutions, thus supporting interdisciplinary discussion and collaboration.

### Missing Voices Impede Effective Governance

The World Health Organization (WHO) has called for dialogue among all stakeholders in the “AI for health ecosystem”, including “developers, manufacturers, regulators, users, and patients.” [5] Yet, our study reveals significant gaps in the voices discussing health-AI’s legal risks. In particular we found an underrepresentation of innovators who build health AI solutions, as evidenced by minimal engagement from authors in computer science and engineering (Figure 4). The apparent lack of engagement is consistent with previous findings of minimal innovator discussion of the legal and ethical dimensions of mental health AI technologies [29]. It is possible that innovators are informed about legal issues but do not actively publish on such matters. However, where innovator engagement is crucial for averting problems, such as through privacy “by design”, our findings are indicative of a concern. Moreover, our findings raise questions about whether heavy regulator reliance on industry voices risks imbalances between innovation enthusiasm and other critical interests like safety and privacy [30].

There is also a notable absence of clinician-driven literature on the complexities of informed consent in AI-assisted treatments. The lack of careful deliberation on this topic risks encouraging blunt solutions. (One public health article suggests, the “[u]se of non-explainable AI should arguably be prohibited in healthcare, where medicolegal and ethical requirements to inform are already high”. [31]) Cross-disciplinary conversations are essential for defining informed consent standards, especially given physicians’ potential lack of training in AI’s risks, and their crucial role in translating medical information for patients. [9–10]

Voices from the global south are also noticeably absent from these discussions, indicating a need for increased inclusion of authors from low- and middle-income countries (LMICs). [32–33] This finding may be partly attributable to our having searched articles published in English and French. However, given known barriers to the full inclusion of academic voices from the Global South, and the disproportionate effects of some legal issues on LMICs (e.g., a heightened risk of biased algorithms perpetuating existing inequalities and compromising patient safety), meaningful engagement from LMIC stakeholders is crucial for realizing the global potential of health AI.

### The Importance of Multidisciplinary Analysis on key Issues

Our analysis reveals divergent disciplinary perspectives on key issues, such as liability and equity, which risk undermining effective AI governance if they are not understood and reconciled. A collaborative approach is essential for ensuring that regulation is fair, appropriately balancing competing interests, perspectives, and concerns, and that it is effective, able to achieve its intended goals.

For instance, to more clearly allocate responsibility when AI leads to patient harm, some jurisdictions give lighter regulatory scrutiny to health innovations where physicians remain involved. [34] This dynamic shifts responsibility to physicians who may not have the information or training needed to evaluate AI processes or outcomes. We need interdisciplinary cooperation to allocate responsibility fairly and build trust. Another key area of concern is the prevention of inequities. There are tensions between incentivizing beneficial innovation through patent protection and ensuring equitable access to technology and data for public interest research and care. Ensuring nuanced, cross-disciplinary analysis, can aid in our understanding and balancing of these competing interests and concerns.

Collaboration between disciplines, with their different expertise and perspectives, will also help ensure that regulation is effective, achieving its intended aims. For example, while many emphasize the need for stronger privacy protections, this need may collide with the need for data, including data relating to race and socioeconomic status, to train algorithms so they are generalizable to different populations. As one author puts it, unless we effectively address biases in AI, “patients that have historically not benefited from the healthcare industry will continue to face discrimination”; our current biases will “become solidified, automated ones.” [35] Interdisciplinary discussion can help us to understand where well-intentioned legal developments (e.g., to strengthen privacy) might have unintended effects (e.g., undermining equity).

Another example is direct-to-consumer health-AI tools like mental health apps, care-robots, and mobility devices. Some argue these are important tools for filling troubling gaps in healthcare service provision. Yet, others observe that insufficient regulatory oversight could undermine that aim and harm vulnerable users (“bots could be programmed to infiltrate people’s homes and lives en masse, befriending children and teens, influencing lonely seniors, or harassing confused individuals until they finally agree to services that they otherwise would not have chosen.” [36–37] These debates underscore the need for cross-disciplinary input, including from those whose lives are affected by health-AI, to achieve equitable and non-discriminatory health AI. [38]

National and international leaders increasingly advocate for interdisciplinary collaboration on AI regulation. [5, 8, 39] Yet, arguably, current regulatory proposals remain unduly siloed. [40] Our study supports calls for more meaningful interdisciplinarity, demonstrating the value of diverse stakeholder input to strike the right balance between competing values, and respond effectively to rising concerns.

## IV. CONCLUSION

Governments must facilitate cross-disciplinary discussions to address legal risks and solutions in health AI effectively. Collaboration across disciplines is essential for guiding the governance of health AI to ensure equitable, safe, and responsible advancements for all.

## Data Availability

All data produced in the present study are available upon reasonable request to the authors

## ACKNOWLEDGEMENTS

We thank Karni A Chagal-Feferkorn, Nicole Davidson, Samantha Iantomasi, Arianne Kent, Kelli White, Caroline Mercer, Angie Ortiz-Romero, and Saly Sadek for assistance, including with research, article review, and data extraction. We also thank the Canadian Institutes of Health Research, the Hospital for Sick Children Research Institute, and the Alex Trebek Forum for Dialogue for funding. The funders had no other role in this study.

## RESEARCH ETHICS APPROVAL STATAEMENT

Research ethics approval was not required for this study as it does not involve human participants.

## DATA SHARING STATEMENT

The data supporting the findings of this study, including a list of all included studies and the extracted data, will be available via a public GitHub link, <https://github.com/mohsensheikhhassani/ScopingReview/tree/main>.

## Online Supplement

**e-Table 1.** Sources of data.

|  |  |  |
| --- | --- | --- |
| **Searched databases** | MEDLINE® (Ovid), EMBASE (Ovid), HeinOnline Law Journal Library, Index to Foreign Legal Periodicals (HeinOnline), Index to Legal Periodicals and Books (EBSCOhost), Web of Science (Core Collection), Scopus, and IEEE Xplore. |  |
| **Databases excluded from searching** | Lexis Advance Quicklaw, WestlawNext Canada | *Rationale:* Significant limitations in the search interfaces of these databases made systematic searching and exporting of results very challenging. Moreover, the vast majority of journals covered by the Lexis and Westlaw databases are also covered by HeinOnline. HeinOnline indexes 4253 law journals, including almost all U.S. law journals in addition to international journals. This coverage was improved further by combination with Gale LegalTrac (1200+ major law reviews) and the H.W. Wilson Index to Legal Periodicals (1100+ legal journals), which also provide coverage for Canada, Great Britain, Ireland, Australia and New Zealand. An analysis by HeinOnline shows coverage of 99/100 of the top US law journals and much more comprehensive coverage of these journals than either Westlaw or Lexis Nexis. [1] |

**e-Table 2.** MEDLINE Search Strategy.

| **e-Table 2. MEDLINE Search Strategy** |  |  |
| --- | --- | --- |
| # | Search | Results |
| 1 | artificial intelligence/ | 26391 |
| 2 | machine learning/ | 16485 |
| 3 | deep learning/ | 6249 |
| 4 | supervised machine learning/ | 931 |
| 5 | unsupervised machine learning/ | 506 |
| 6 | natural language processing/ | 4799 |
| 7 | neural networks, computer/ | 31313 |
| 8 | robotics/ | 22242 |
| 9 | ((machin* or artific* or comput* or robot* or automat*) adj3 intelligen*).ti,ab,kf. | 16925 |
| 10 | ((assist* or augment* or autonomous) adj1 intelligen*).ti,ab,kf. | 353 |
| 11 | ((machin* or deep or transfer or hierarchical) adj2 learning).ti,ab,kf. | 65348 |
| 12 | (autoML or robot* or droid* or android* or telerobot* or tele-robot* or (remote* adj2 operat*)).ti,ab,kf. | 57138 |
| 13 | natural language processing.ti,ab,kf. | 4541 |
| 14 | neural network*.ti,ab,kf. | 62994 |
| 15 | (comput* adj2 (reason* or vision* or knowledg* or cogniti*)).ti,ab,kf. | 8819 |
| 16 | (perceptron* or connectionis*).ti,ab,kf. | 4215 |
| 17 | legislation as topic/ | 15972 |
| 18 | legislation, hospital/ | 2455 |
| 19 | legislation, medical/ | 16581 |
| 20 | medical device legislation/ | 271 |
| 21 | international health regulations/ | 66 |
| 22 | legislation, nursing/ | 3161 |
| 23 | legislation, pharmacy/ | 1266 |
| 24 | privacy/ | 6934 |
| 25 | jurisprudence/ | 29909 |
| 26 | confidentiality/ | 23828 |
| 27 | contracts/ | 3304 |
| 28 | informed consent/ | 37305 |
| 29 | informed consent by minors/ | 231 |
| 30 | third-party consent/ | 3776 |
| 31 | parental consent/ | 3298 |
| 32 | intellectual property/ | 1602 |
| 33 | patents as topic/ | 10109 |
| 34 | copyright/ | 682 |
| 35 | international law/ | 100 |
| 36 | legal services/ | 37 |
| 37 | malpractice/ | 28036 |
| 38 | liability, legal/ | 15815 |
| 39 | ownership/ | 9226 |
| 40 | (law* or legislat* or legal* or medico-legal or medicolegal or statut* or bylaw or by-law* or court* or litigat* or juris* or constitution* or contract or contracts or contractual*).ti,ab,kf. | 359878 |
| 41 | (confidentiality or (confidential adj3 information)).ti,ab,kf. | 12509 |
| 42 | (privacy adj2 (data or genetic* or patient* or health)).ti,ab,kf. | 3523 |
| 43 | (consent* adj2 (third-part* or informed or minor* or parent* or spous* or communit*)).ti,ab,kf. | 43919 |
| 44 | (intellectual propert* or patent* or trade secret* or (propert* adj2 right*)).ti,ab,kf. | 58492 |
| 45 | copyright*.ti,kf. | 368 |
| 46 | (liability or liabilities or tort or torts or tortious or malpractice or negligen*).ti,ab,kf. | 35518 |
| 47 | (treaty or treaties).ti,ab,kf. | 2011 |
| 48 | human rights/ | 14839 |
| 49 | civil rights/ | 10086 |
| 50 | patient rights/ | 7150 |
| 51 | right to health/ | 147 |
| 52 | (right* adj2 (civil or human or patient* or health or healthcare or minorit* or equal* or collective)).ti,ab,kf. | 33791 |
| 53 | government regulation/ | 21607 |
| 54 | (oversight adj2 government*).ti,ab,kf. | 134 |
| 55 | (regulat* adj3 (government* or federal* or provincial* or state or oversight or requirement* or framework* or guideline* or authorit* or agenc* or body or bodies or data or device* or health or healthcare or medical or approval* or compliance or hurdle* or obstacle* or barrier* or issue*)).ti,ab,kf. | 71321 |
| 56 | (regulations or regulatory environment*).ti,ab,kf. | 49909 |
| 57 | (guidances or guidance document*).ti,ab,kf. | 2188 |
| 58 | or/1-16 | 215779 |
| 59 | or/17-57 | 711869 |
| 60 | 58 and 59 | 5126 |
| 61 | limit 60 to yr="2012 -Current" | 3918 |

**e-Table 3.** Key Terms Defined for Eligibility Assessment.

| **e-Table 3. Key Terms Defined for Eligibility Assessment** |  |
| --- | --- |
| **Legal Concerns** | Law is “the formal rules of a country passed by a government or its delegated representatives to regulate conduct”. [2,3]<br>This encompasses formal laws (e.g., constitutions, statutes, the common law) and regulations (e.g., rules passed pursuant to statutory authority).<br>‘Law’ here does not include ‘soft’ law (e.g., professional college rules).<br>A ‘legal concern’ is one that is identified as requiring a formal governmental response. |
| **Artificial Intelligence (AI)** | Per the World Health Organization, ‘artificial intelligence’ (AI) is “the ability of algorithms encoded in technology to learn from data so that they can perform automated tasks without every step in the process having to be programmed explicitly by a human” and as the “performance by computer programs of tasks that are commonly associated with intelligent beings.” [4]<br>AI, then, refers to machines that can perform acts that typically require human cognition without direct human assistance. This covers a range of tools including those that read medical images to possible future surgical robots.<br>AI does not include electronic tools that merely aid in data collection that do not have an associated AI component (e.g., wearable sensors, computer-assisted decision supports). |
| **Health-Related** | AI in this review is ‘health-related’ if it pertains to “healthcare” or “public health.”<br>Healthcare is understood as “efforts made to maintain or restore physical, mental, or emotional well-being especially by trained and licensed professionals”. [5]<br>This also includes activities by trainees or AI where the trainee or AI functions in the same capacity as the licensed professional.<br>“Public health” is understood as “the art and science dealing with the protection and improvement of community health by organized community effort and including preventive medicine”. [6]<br>This definition focuses on activities typically performed by health professionals (and those serving their functions) and the organization of the healthcare settings in which they work.<br>It includes activities in and the regulation of hospital, physician, long-term care home, and other healthcare provider settings as well as at-home goods and services for curative, diagnostic, and preventative purposes.<br>For greater certainty, it includes activities performed by healthcare professionals and basic features of healthcare systems and their regulation (e.g., rationing decisions, insurance). |

**e-Table 4:** Key Variables for Data Extraction.

| Possible Author Disciplines (faculty of corresponding author) | Possible Legal Issues (must be discussed “as a legal issue”) | Possible Solutions |
| --- | --- | --- |
| Law | Privacy / data protection | New Legislation or Regulations (E.g., A New Legislative Framework for Health-Related A.I.) |
| Medicine | Safety or quality | Reform of Existing Laws or Regulations (E.g., Amendments to Existing Privacy Laws) |
| Nursing | Bias or discrimination | Professional Guidelines Issued by a Body with Regulatory Power (E.g., the College of Physicians and Surgeons or Professional Engineers Ontario, the F.D.A., Health Canada) |
| Pharmacy | Liability | Non-Binding Guidelines Issued By Other Bodies Without Regulatory Powers (E.g., The World Health Organization) |
| Other health professions (Dentistry, nutrition, etc.) | Informed consent | Mandatory Professional Training (As Part of Degree or Licencing Requirements or Continuing Education) |
| Public Health | Legal capacity | Calls for Voluntary Improvements (By the Industry as a Whole, Individual Firms, Innovators, Providers, Etc.) |
| Computer science | Intellectual property | Other (please list) |
| Engineering | Efficiency of regulation (e.g., over-/under-regulation, the impact of regulation on innovation, discussed as a general matter) |  |
| Other (please specify) | Legal personhood |  |
|  | Access and Insurance |  |
|  | Other legal issue |  |

**e-Table 5:**
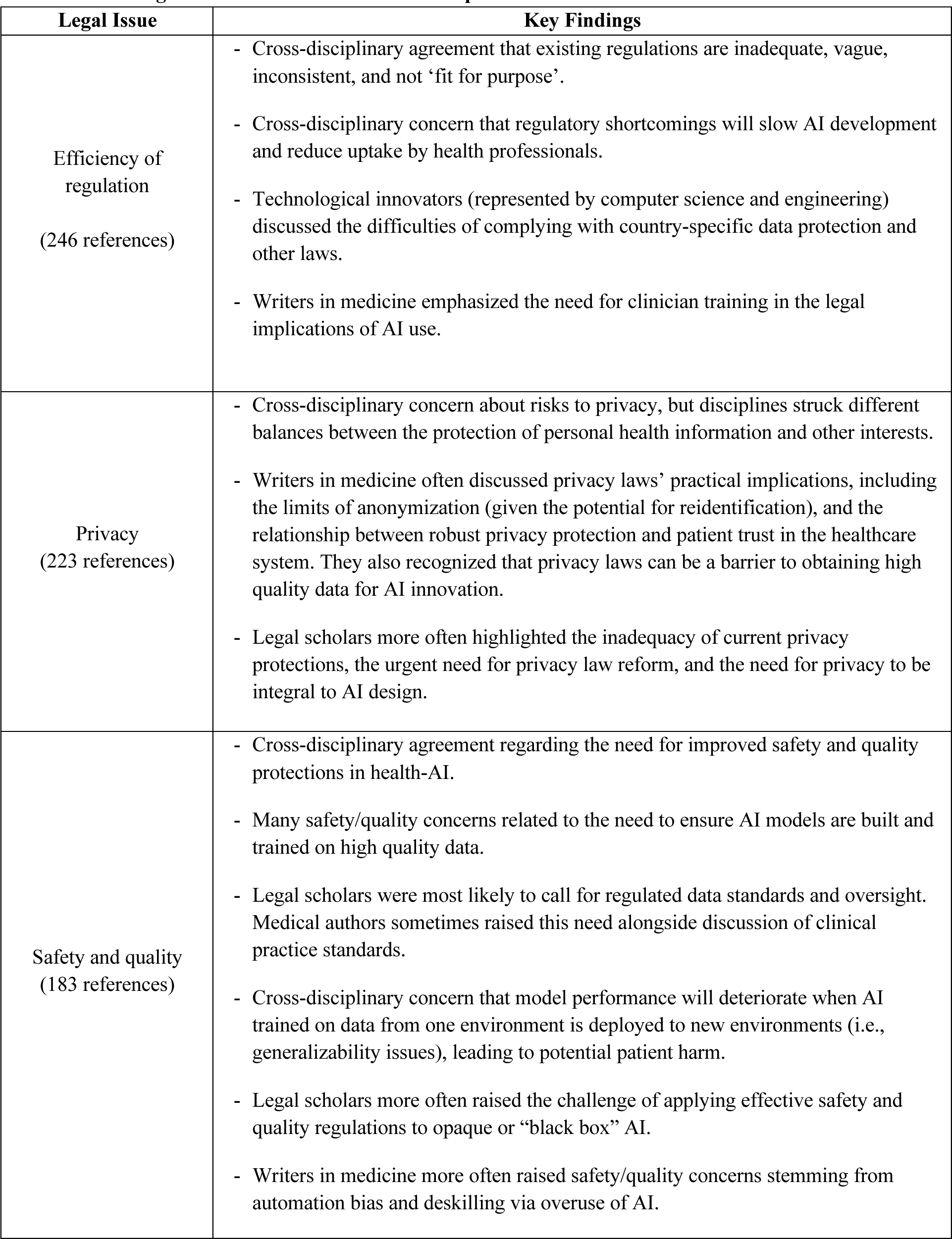
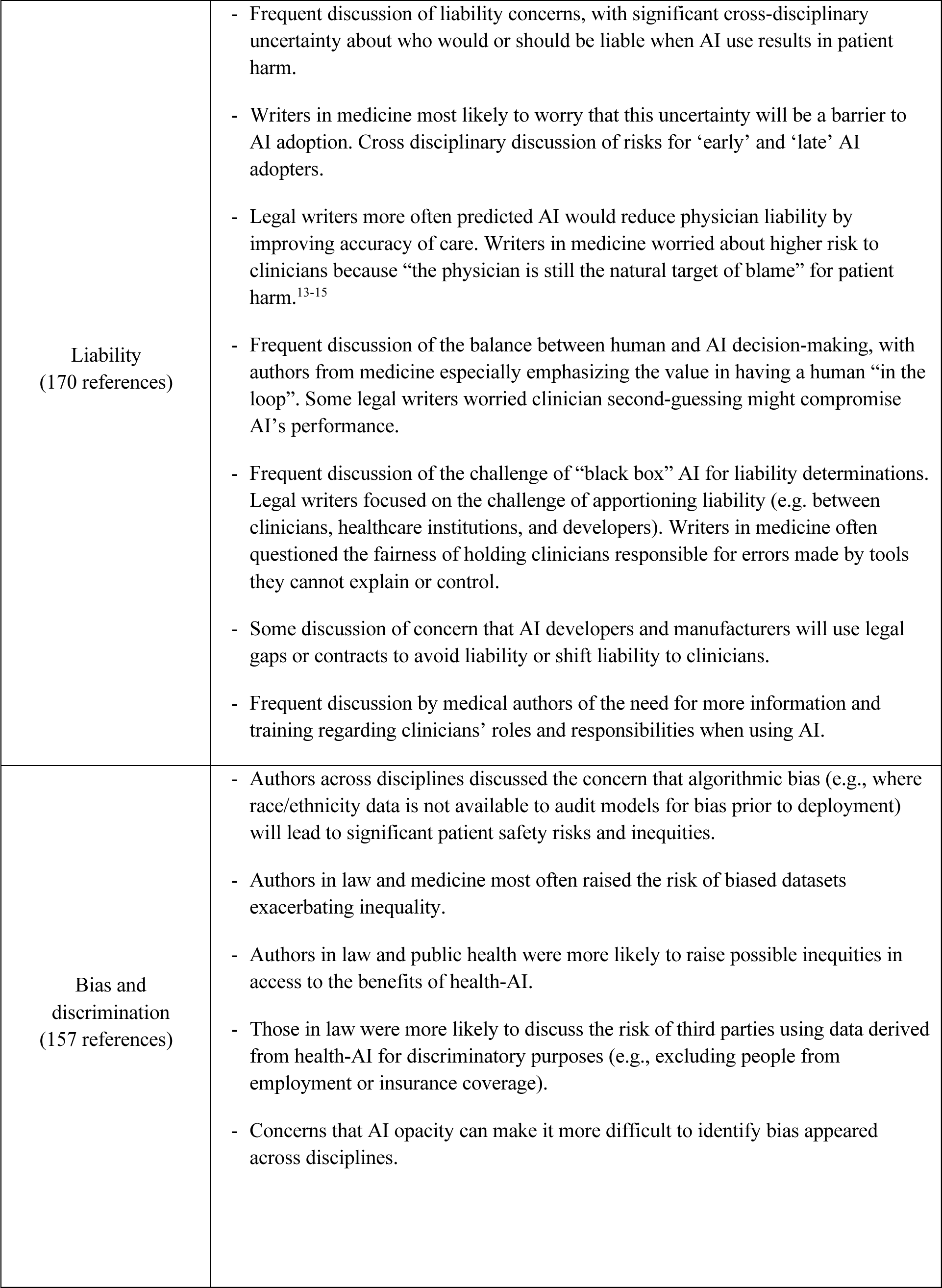
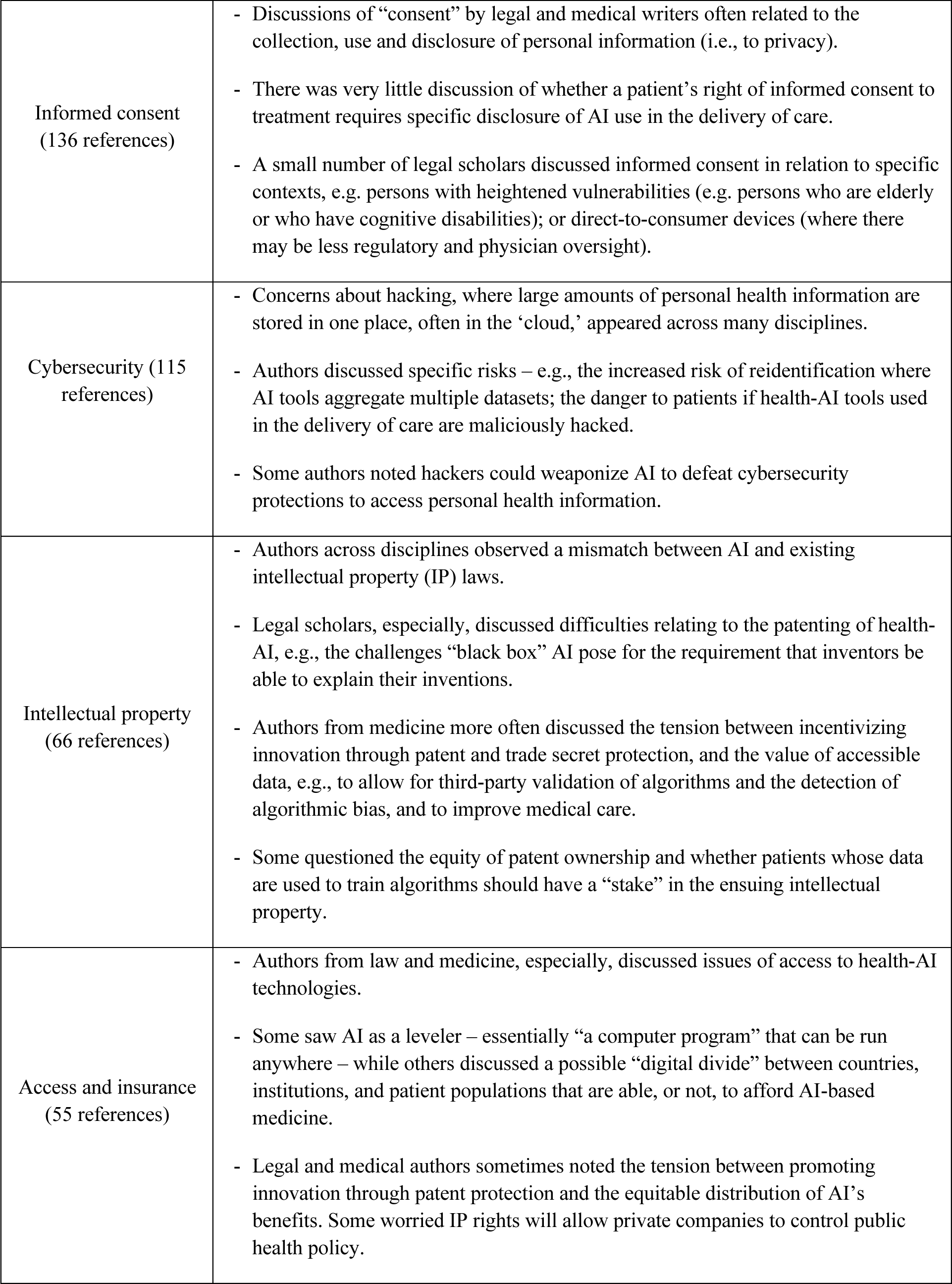
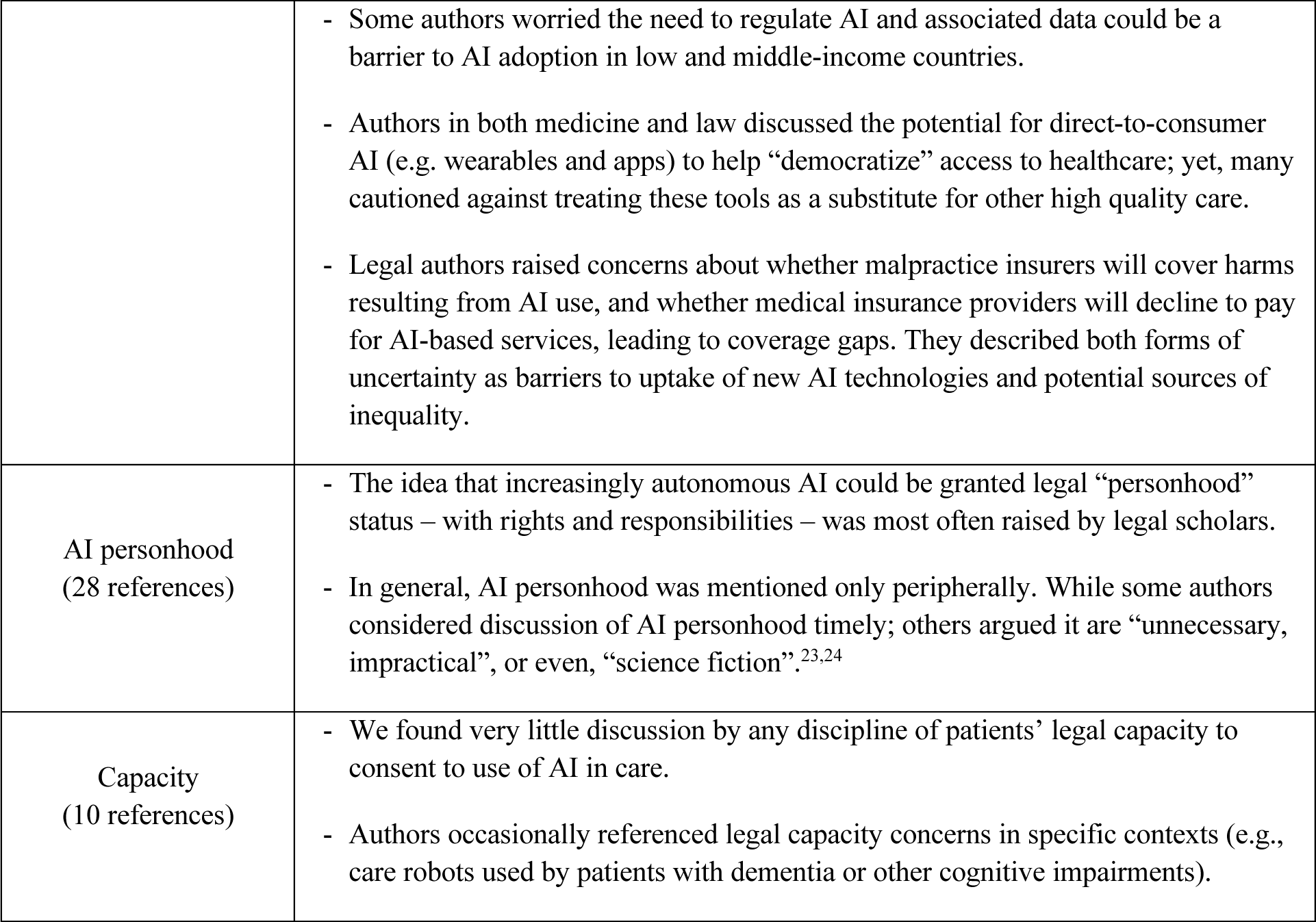
How legal issues are discussed across disciplines.

